# Development and validation of a risk prediction algorithm for high-risk populations combining genetic and conventional risk factors of cardiovascular disease

**DOI:** 10.1101/2025.04.02.25324383

**Authors:** Tuuli Puusepp, Ave Põld, Lili Milani, Aet Elken, Estonian Biobank Research Team, Mikk Jürisson, Krista Fischer

**Author notes:** **Corresponding author:** Tuuli Puusepp.

## Abstract

**Aim:** To develop a model for cardiovascular disease (CVD) risk, combining polygenic risk score (PRS) with traditional risk factors while assessing the added value of PRS in two cohorts of biobank participants.

**Methods:** Data of 128 209 participants from the Estonian Biobank recruited between 2003– 2011 and 2018–2019 without prevalent cardiovascular disease, was included. Hazard ratios (HR) for polygenic risk versus conventional risk factors were estimated with Cox proportional hazards models, cumulative incidence was assessed with Aalen-Johansen curves. Predictive performance was tested using a split-sample approach and competing risk modelling. Age at CVD event served as the outcome, and the impact of the PRS was evaluated by age group (25–59 vs. 60+), sex, and recruitment period, using HRs, Harrell’s C-index, and net reclassification indices (NRI).

**Results:** The estimated HR per one standard deviation (SD) of PRS ranged from 1.1, 95% CI 1.06–1.15 (age 60+, earlier cohort) to 1.36, 95% CI 1.24–1.49 (men 25–59, later cohort). Adding PRS to the conventional risk factors in the age group 25–59 increased the C-statistic by 0.028 (p<0.0001) for men. In the age group 60+, the increase was 0.016 (p=0.0002) across all. In the independent validation set, the continuous NRI was 19.1% (95% CI 13.3%–24.9%) in the 25–59 group and 13.9% (95% CI 8.1%–19.6%) in the 60+ group.

**Conclusions:** In a high-risk population, PRS is a strong independent risk factor for CVD and should be considered in routine risk assessment, starting at a relatively young age.

## Introduction

Atherosclerotic cardiovascular diseases (CVD), including coronary artery disease and cerebrovascular disease, are the leading cause of death in numerous European countries [1]. Polygenic risk scores (PRSs) have demonstrated their effectiveness as a valuable and innovative method for assessing genetic risk related to CVD and improving the accuracy of disease risk prediction[2]. Several studies indicate that incorporating genetic risk assessment into existing risk stratification algorithms could significantly enhance their efficiency [3], [4], [5].

Polygenic risk scores have been validated in several studies and have been found to enhance CVD risk prediction independently of many traditional factors such as smoking, hypercholesterolemia, hypertension, obesity, and family history of CVD [2], [6], [7] [8]. Typically, a PRS combines the effect of a large number (from hundreds to millions) of single nucleotide polymorphisms (SNPs) as a weighted sum of allele counts [9]. It has been shown that elevated polygenic scores contribute to a significantly higher percentage of early-onset myocardial infarction cases than monogenic variants for familial hypercholesterolemia [10], [11]. This implies that integrating genetic predisposition complements CVD risk prediction and, when combined with traditional factors, can significantly improve disease risk prediction and facilitate decision making in primary prevention of CVD [12]. Studies assessing CVD risk combining PRS with clinical and lifestyle data show promising results, yet more rigorous validation and comparisons between existing models are necessary to justify their clinical utility [12], [13], [14], [15], [16].

Several large cohort and country-specific prognostic CVD risk models have been developed based on traditional risk factors such as the European SCORE2, the American pooled cohort equations (PCE), and the UK-specific QRISK3 algorithm, yet their generalisability across different populations remains limited [17], [18], [19], [20], [21]. A comparable challenge arises when considering the questionable effectiveness of PRS-based risk assessment algorithms that utilize UK Biobank data, given that the United Kingdom has a low prevalence of cardiovascular disease compared to high-risk populations in Middle and Eastern Europe [22], [23], [24].

This study describes the development and validation of the novel risk assessment model combining traditional cardiovascular risk factors with PRSs.

## METHODS

### Sources of data

Data from the Estonian Biobank (EstBB) was used to compose the study cohort [25]. EstBB is a volunteer-based biobank that includes genotype and clinical events data of more than 210 000 participants. The health records are regularly updated using national registries, hospital databases and the database of the national health insurance fund which covers data from both primary and secondary care. Additionally, the cholesterol data was quantified using nuclear magnetic resonance (NMR) spectroscopy. Participants have been recruited during two distinct phases: 1) an earlier cohort (2002–2017) including 52 266 participants mainly recruited by general practitioners and 2) a later cohort (2018–2022) including 159 102 participants that joined the biobank during a national campaign [25].

The PRS used in this study is the multi-ancestry PRS for coronary artery disease developed by Patel, et al. and obtained from the PGS catalog [26], [27]. This PRS was selected from a pool of 151 candidate CAD PRSs for its highest z-score in predicting prevalent CVD via logistic regression adjusted for age at recruitment and sex, utilising EstBB data comprising 15 095 CVD cases and 119 694 controls at baseline.

### Participants

#### Sample size

All 185 760 EstBB participants aged at least 25 years at recruitment with genotyping data available were considered for the analysis. After applying the inclusion and exclusion criteria (see below), the total number of individuals included in the study was n=128 209.

#### Exclusion criteria

– Prevalent CVD cases i.e. individuals diagnosed with non-fatal CVD (ICD-10 codes I20, I21–I25, I60–I69 excluding I60, I62, I67.1, I67.5, I68.2) before recruitment (n=25 894)
– Individuals with diabetes mellitus (E10–E14) at baseline (n=11 555)
– Individuals with familial hypercholesterolemia (FH) (n=76)
– Individuals with missing lipid values (Total cholesterol (Total-C), HDL cholesterol (HDL-C)) (n=3 569)
– Individuals with missing systolic blood pressure (SBP) or with SBP<50 mmHg or SBP>300 mmHg (n=27 212)
– Individuals with missing body mass index (BMI) or with BMI less than 15 kg/m2 or more than 50 kg/m2 (n=752)
– Individuals with missing smoking data (n=1 591)

#### Outcome

The outcome of interest was incident non-fatal or fatal CVD event. Incident CVD events were identified using the atherosclerotic CVD definition provided by the SCORE2 working group [17]. A detailed list of the ICD-codes is in Table S1. As the outcome data is based on Electronic Health Records (EHR) linkage, we assumed there is no missing outcome data. The data from EHR was available up to December 31^st^, 2023. The outcome event was observed in 6 893 individuals and deaths from non-CVD causes (n=2 124) were treated as competing events.

#### Predictors

The model combined conventional predictors of CVD and pre-calculated PRS for CAD. Predictor management has been described in the Supplementary file.

List of the included predictors:

– Age (years)
– Sex (M/F)
– Current smoking (y/n)
– SBP (mmHg)
– Total cholesterol (mmol/L)
– HDL cholesterol (mmol/L)
– BMI (kg/m2)
– PRS for CAD

#### Statistical methods

a) Impact of PRS on the outcome Aalen-Johansen curves, accounting for competing causes of death and using age as time scale, were estimated to assess how PRS differences impact cumulative CVD event incidence in men and women aged 25–70 at recruitment across three PRS groups (bottom 10%, 10%– 90%, top 10%). Crude hazard ratios with 95% CIs were calculated using the Cox proportional hazards models, separately for the earlier and later cohort.
b) PRS effect and discrimination compared with that of conventional risk factors Separate models were fitted for earlier and later cohort and two age groups (25–59 and 60+ years) to estimate the effect of PRS and compare its discrimination with that of conventional risk factors (current smoking, SBP, total cholesterol, HDL cholesterol, BMI). In the younger group, models were sex-specific due to significant interactions, while in the older group, sex was a stratification variable. Cox models used age at CVD event as the time scale to avoid bias due to left truncation. Harrel’s C-index was used to assess discrimination for single-risk-factor models and models with all covariates, with and without PRS.
c) Model’s predictive ability in an independent sample

An independent dataset was used to assess the model’s predictive ability using a split-sample approach, dividing the cohort into a training set (one-third) and a validation set (two-thirds of the cohort). In each group (defined by age, sex, and recruitment period), two Cox proportional hazard models were fitted in the training datasets with time from recruitment to the incident CVD event as the main outcome of interest. The models accounted for competing risks using the multi-state modelling principle, where other causes of death were considered as competing events [28].

The event times for participants with no events or with an event occurring later than 5 or 10 years after recruitment were censored at 5 or 10 years for the later and the earlier cohort, respectively. Models were first fitted using traditional CVD risk factors, then with these factors plus PRS.

Model calibration was assessed by comparing the number of observed CVD events within quintiles of predicted risk with those predicted from the models. Continuous net reclassification index (NRI) and categorical NRI were computed to compare the 5-year predicted risk with observed event rates. To convert the linear predictors from the survival models adjusted for traditional risk factors and the PRS into absolute 5– or 10-year CVD risk predictions, the absolute risk formula of Benichou and Gail was used (implemented in the *riskRegression* package version 2023.9.20 of the R software) [29], [30]. All analyses were done using R version 4.2.2 [31].

## RESULTS

### Cohort description and outcomes

A total of 128 209 Estonian Biobank participants were included in the analysis. Baseline characteristics of the earlier cohort (n=32 554, recruited in 2002–2017) and later cohort (n=95 655, recruited in 2018–2022) are presented in Table 1. The mean age at recruitment in both cohorts is similar, approximately 44 years (44.4 in the earlier cohort and 43.7 in the later) and the sex distributions are comparable (about two thirds of the cohorts being women). The median follow-up period is 14.9 years for the earlier cohort and 5.1 years for the later cohort. Regarding risk factors, there is a large difference in smoking prevalence: 30% of the earlier cohort were current smokers at recruitment, compared to 19% in the later cohort. We can also see somewhat higher average blood pressure and LDL cholesterol levels in the earlier cohort among individuals aged 60 and older. The 5-year cumulative CVD incidence differs being 5.7% in the earlier and 0.6% the later cohort.

**Table 1.**
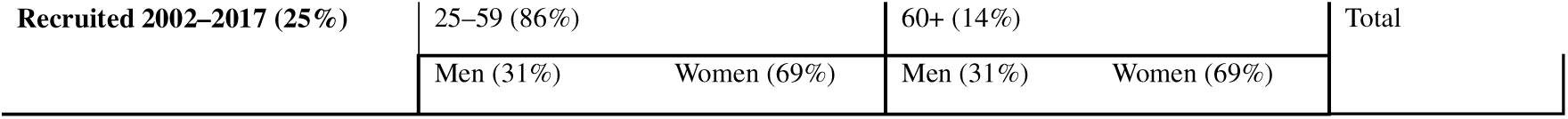
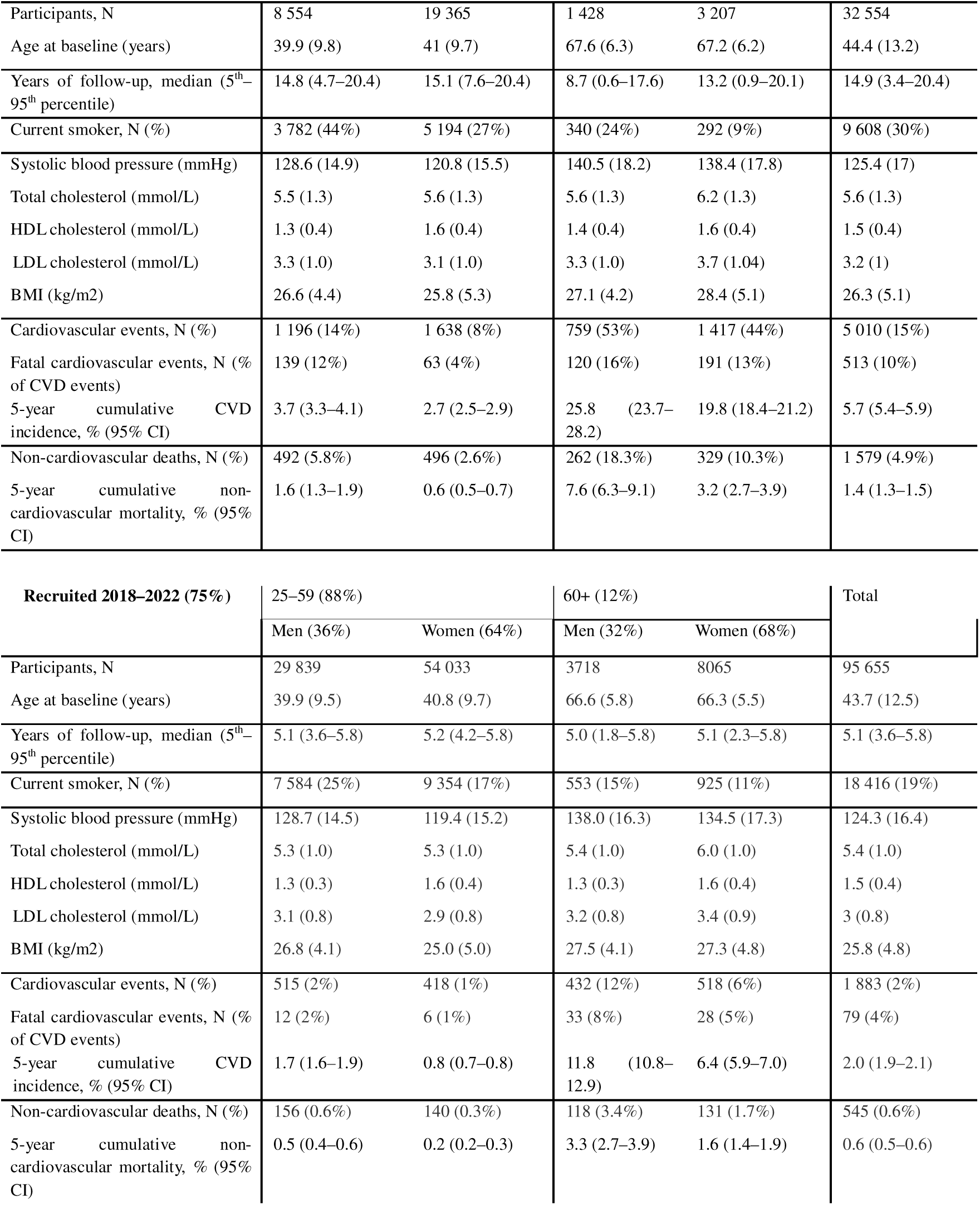
Baseline participant characteristics. Data are mean (SD) unless noted otherwise.

### Cumulative incidence in different PRS percentiles

The cumulative incidence curves on age scale (Figure 1) show higher CVD incidence in men than women across all PRS groups and cohorts. By age 70, about 40% of men in the earlier cohort had experienced CVD, compared to about 25% in the later cohort. For women, the rates were 24% and 13%, respectively. Men in the later cohort had a similar CVD risk to women in the earlier cohort.

**Figure 1.**
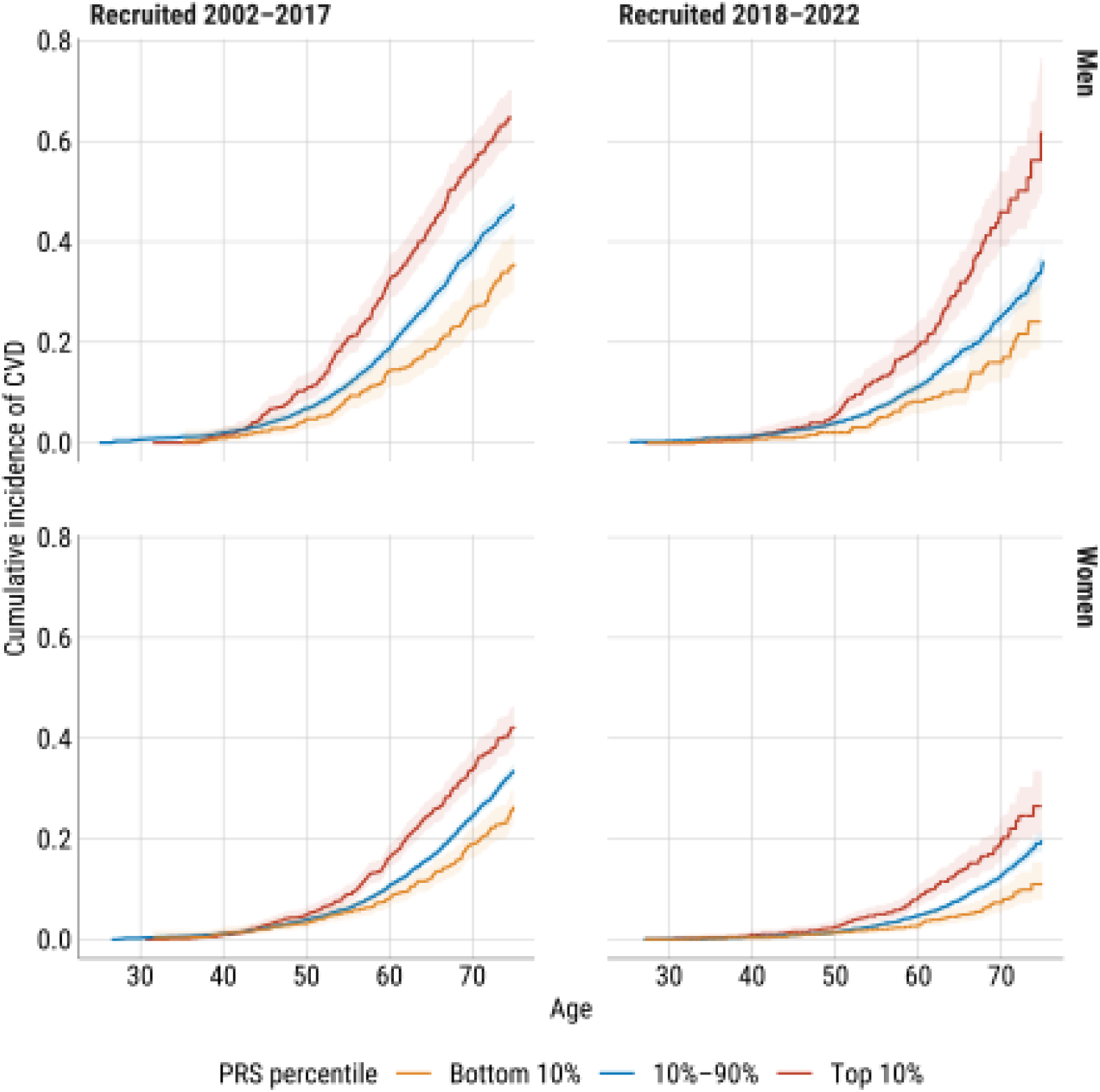
Cumulative incidence of CVD with 95% CI in men and women aged 25–70 at recruitment by PRS percentiles and recruitment period using age as time scale.

The effect of PRS is consistent across sexes and cohorts. Individuals in the highest PRS decile have significantly higher CVD risk compared to those in the middle or lowest percentiles. For example, in the earlier cohort, men in the highest PRS decile have over double the cumulative incidence of CVD by age 70 compared to those in the lowest decile. Men in the top PRS decile reach a 20% cumulative incidence of CVD six years earlier than average, while those in the lowest decile reach it five years later, creating a decade-long difference between extreme deciles. Similar patterns are observed in women.

Compared to individuals in the 10th–90th PRS percentile range, the hazard ratio (HR) for those in the highest PRS decile is 1.7 (95% CI 1.5–1.9) for men and 1.5 (95% CI 1.3–1.7) for women in the earlier cohort, 1.9 (95% CI 1.6–2.4) for men and 1.6 (95% CI 1.3–2.0) for women in the later cohort. When the highest PRS decile is compared to the lowest, HRs range from 1.9 for women in the earlier cohort to 2.9 for men in the later cohort.

### Effect of PRS in a model with conventional risk factors

HRs corresponding to PRS are shown in Figure 2. These were obtained from Cox models with age as time scale, adjusted for current smoking, SBP, total cholesterol, HDL cholesterol and BMI. Models were fitted separately for the two cohorts, sexes, and age groups (in individuals aged 60+, sex-stratified models were fitted).

**Figure 2.**
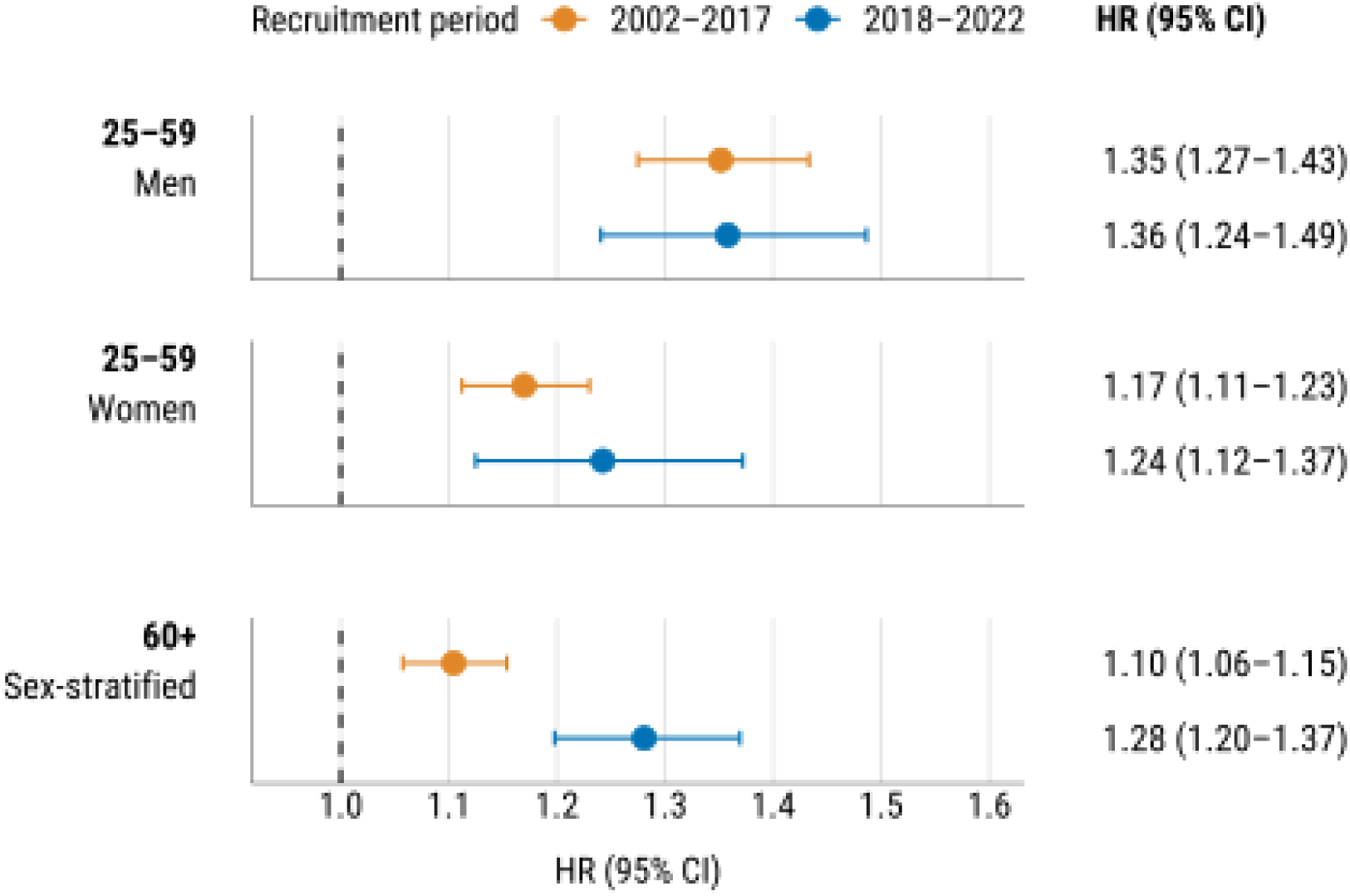
HRs (95% CI) for one SD of PRS by sex, age group and recruitment period from Cox models fitted on the entire data using age as time scale and adjusted for all conventional risk factors.

The effect of PRS is strongest in younger men, with similar HRs across cohorts. Younger women show slightly lower HRs, with a stronger effect in the later cohort. In individuals aged 60+, the effect is more pronounced in the later cohort, with no significant interaction between PRS and sex in this age group. Detailed parameter estimates are shown in Table S2.

### Discriminatory power of PRS and conventional risk factors: comparison of C-indices

Cox models using age as time scale and stratified by cohort (and additionally by sex in the age group 60+) were fitted with each traditional risk factor and PRS as a single covariate to assess their relative importance. Harrell’s C-index estimates remained below 0.6 (Figure 3), reflecting the model’s discriminative ability for individuals of the same baseline age, as age was used as time scale.

**Figure 3.**
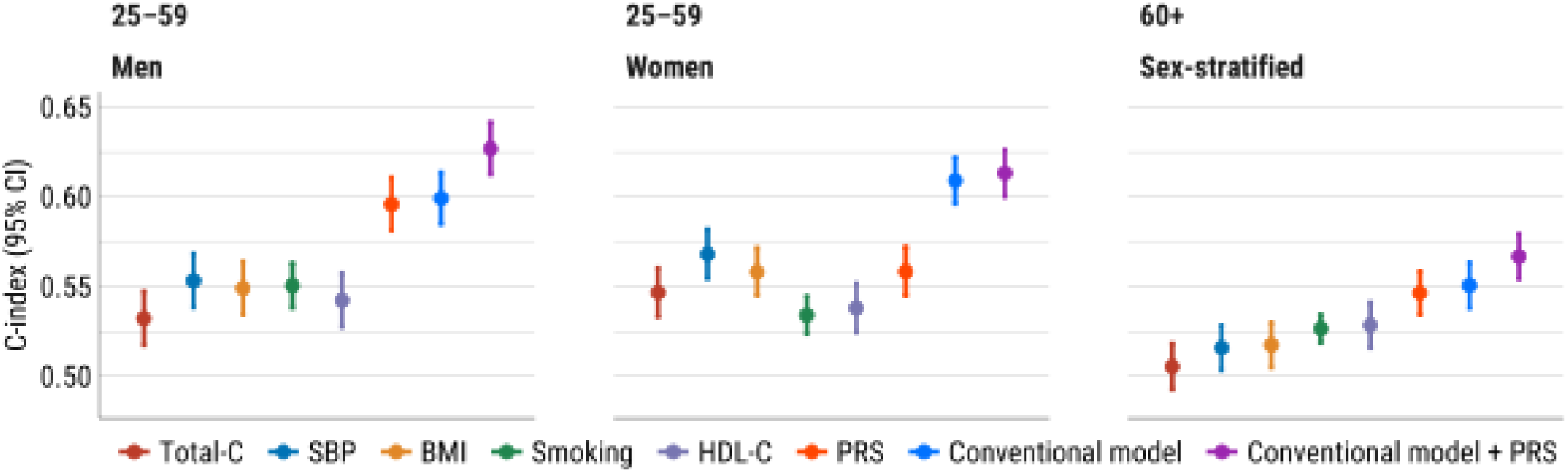
C-indices for individual risk factors, the conventional model, and the PRS added to the conventional model.

In younger men, PRS is the strongest single predictor, with a C-index of 0.60 (95% CI 0.58– 0.61), nearly equivalent to that of the model with traditional risk factors. Combining traditional risk factors and PRS leads to a C-index of 0.63 (95% CI 0.61–0.64), representing an improvement of 0.028 (p<0.0001) over the model without PRS.

In younger women, the effect of PRS alone (C-index 0.56, 95% CI 0.54–0.57) is not stronger than that of SBP or BMI, but it still leads to a slight improvement in the combined risk factor model (C-index 0.61, 95% CI 0.60–0.63, increase 0.004, p=0.085).

In individuals aged 60+, PRS is the strongest single predictor (C-index 0.55, 95% CI 0.53– 0.56). Adding PRS to the traditional risk factor model increases the C-index to 0.57 (95% CI 0.55–0.58, increase 0.016, p=0.0002).

In a separate analysis of the two cohorts, there were only minor differences in C-index values across cohorts, with the exception of 60+ age category, where the improvement was clearly higher in the new cohort (adding PRS increased the C-index by 0.008 in the 2002–2017 cohort and by 0.03 in the 2018–2022 cohort; details in Table S3).

### The performance of model-based predictions in an independent sample

To assess the performance of the predictive algorithm on independent data, models including all traditional risk factors, with and without PRS, were refitted using the training set of 42 827 individuals. The parameter estimates were used to calculate the linear predictor values for 85 382 individuals in the validation set. As seen in Figure S1, calibration of the model with PRS is adequate in the validation set when the predicted risk of CVD in groups defined by quintiles of predicted risk is compared to the number of CVD events in these groups.

Net reclassification analysis comparing the conventional model to the conventional model with PRS were done separately for the two age groups (25–59 and 60+) but jointly for the two cohorts (Table 2). NRI was calculated according to predicted 5-year risk of CVD. For the categorical NRI analyses, risk categories (low, intermediate, and high) were defined as <1.25%, 1.25% to 5%, and >5% within 5 years for the younger age group (25–59), and as <5%, 5% to 10%, and >10% within 5 years for the older age group (60+) [32]. A detailed description of the NRI analysis is in the Supplementary file.

**Table 2.** Results of NRI analysis.

| Age group | 25–59 |  | 60+ |  |
| --- | --- | --- | --- | --- |
| Events/non-events | 1154 | 72858 | 1319 | 9389 |
|  | Value (95% CI) | P | Value (95% CI) | P |
| NRI (%) | 19.1 (13.3–24.9) | <0.0001 | 13.9 (8.1–19.6) | <0.0001 |
| NRI for events (%) | 9.7 (4.0–15.4) | 0.0009 | 7.7 (2.3–13.0) | 0.005 |
| NRI for non-events (%) | 9.4 (8.7–10.1) | <0.0001 | 6.2 (4.2–8.2) | <0.0001 |
| Categorical NRI (%) | 3.0 (1.2–4.8) | 0.001 | 3.1 (1.1–5.0) | 0.001 |

There is a significant improvement in reclassification for both events and non-events in both age groups.

In the age group 25–59, the overall NRI was 19.1% (95% CI 13.3%–24.9%). The categorical NRI in this group showed a modest but significant improvement of 3.0% (95% CI 1.2%– 4.8%).

In the age group 60+, the overall NRI was slightly lower but still significant at 13.9% (95% CI 8.1%–19.6%), and the categorical NRI in this age group was slightly higher at 3.1% (95% CI 1.1%–5.0%). Detailed results of categorical NRI analysis are in Table S4.

Figure 4 displays results of the categorical NRI analysis, focusing on the reclassification of individuals initially classified in the intermediate risk group. In the age group 25–59, the initial intermediate risk category included 19 871 individuals, with a 5-year CVD incidence of 2.6%. After PRS adjustment, 8.1% of these individuals were reclassified to low risk, where the incidence was 0.9%, and 3.7% were moved to high risk, where the incidence increased to 5.8%. In the age group 60+, the initial intermediate risk category included 2 931 individuals, with a 5-year CVD incidence of 8.8%. After PRS incorporation, 15.1% were reclassified to low risk, where the incidence was 5.4%, and 11.4% to high risk, where the incidence was 12.9%.

**Figure 4.**
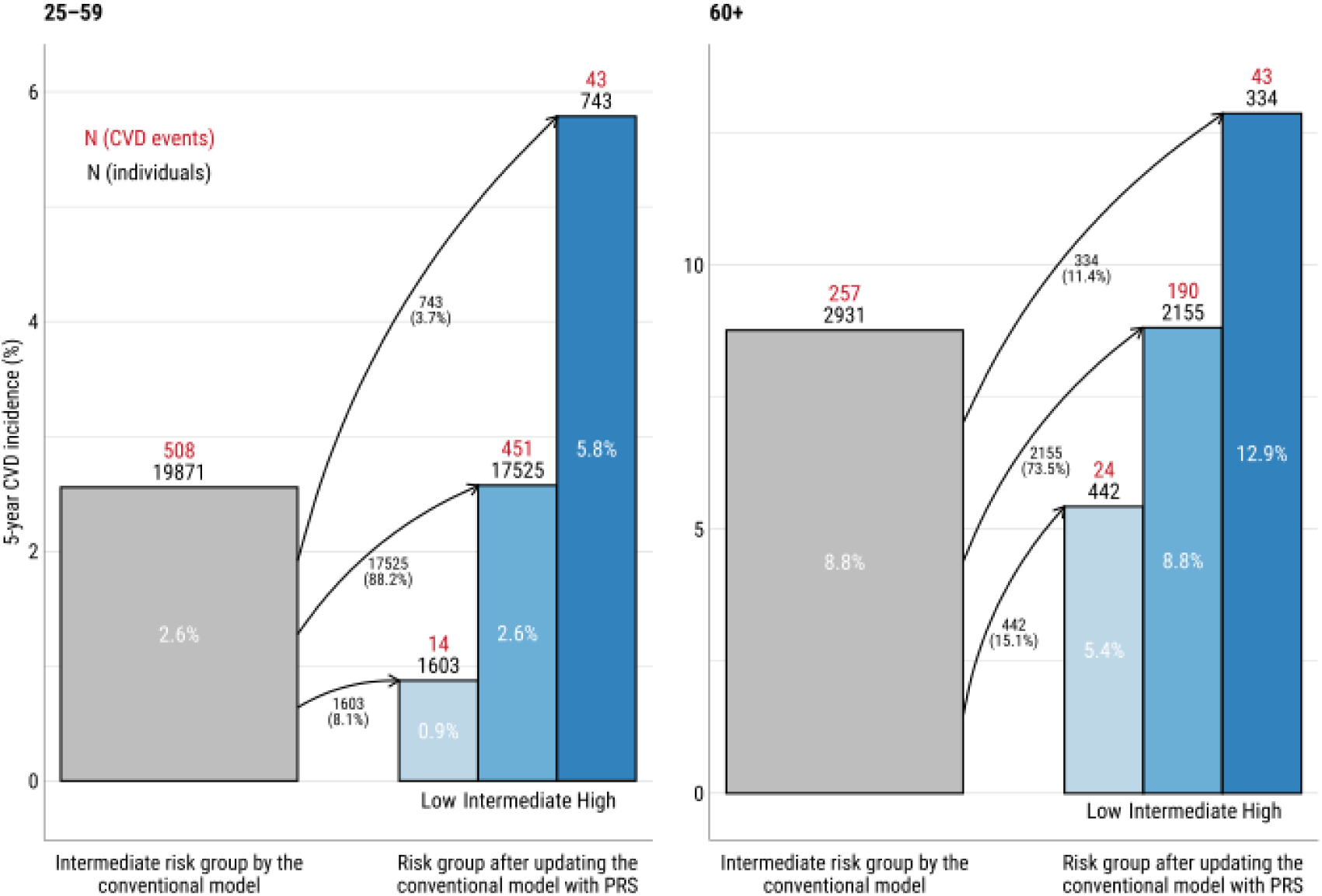
Reclassification of individuals initially categorized as intermediate risk for 5-year CVD incidence using the conventional model. Arrows indicate the movement of individuals between categories, with corresponding percentages representing the proportion of individuals reclassified.

### Potential for using PRS-based risk prediction in practice: an illustration

The practical use of the proposed risk prediction algorithm involves communication of the effect of unmodifiable genetic component (PRS) alongside potentially modifiable risk factors. We illustrate the message that could be delivered to men and women of age 50. Figure 5 shows the predicted 10-year risk for non-smoking individuals with average PRS, as well as predictions for individuals whose PRS exceeds the mean by two SDs and/or who are current smokers. The values of all other risk factors were fixed at their mean values for men or women aged 25–59 in the 2002–2017 cohort.

**Figure 5.**
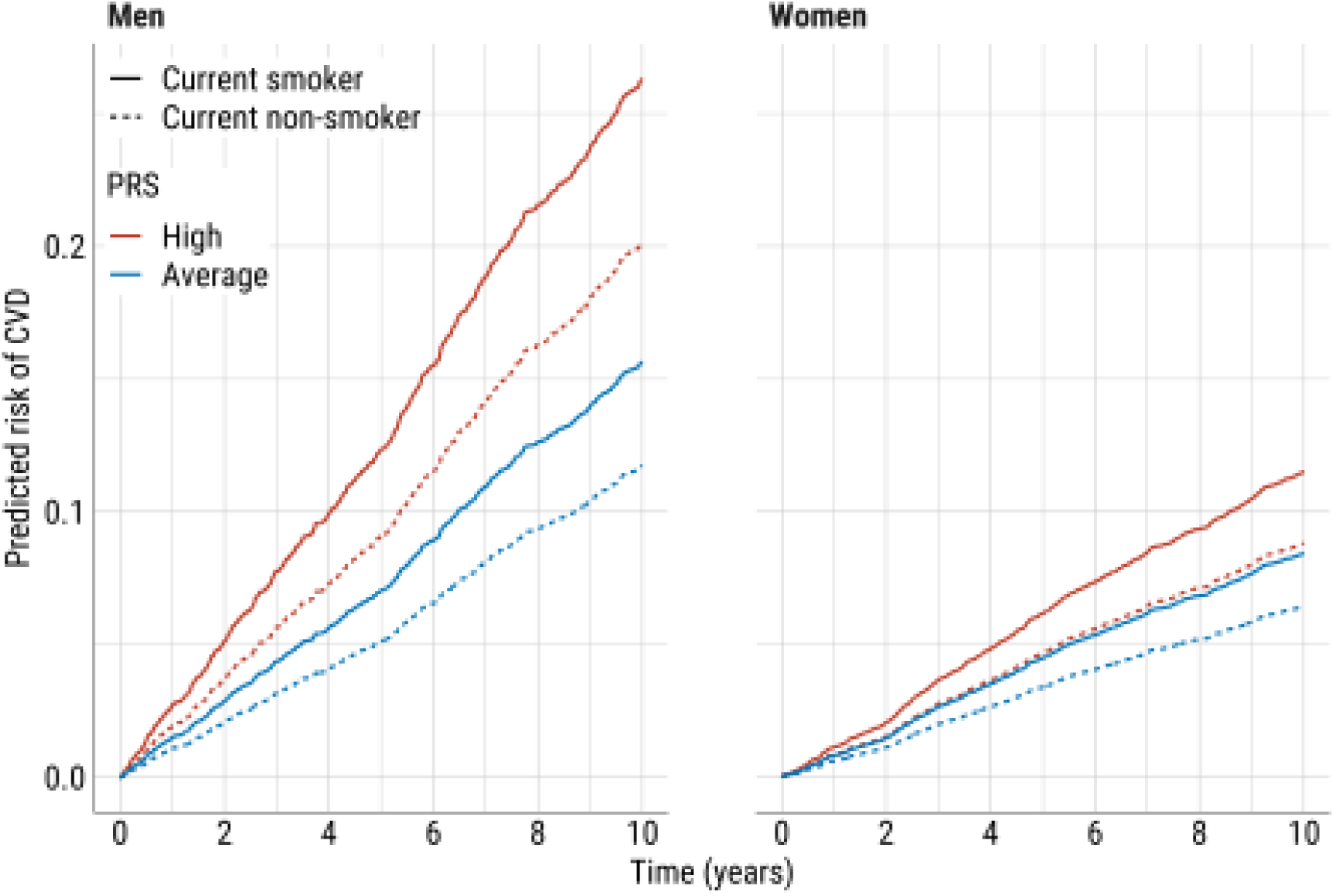
Predicted risk of CVD for men and women aged 50 at recruitment by PRS value and current smoking status. The predictions are calculated from the Cox model fitted using the full data of the age group 25–59 recruited in 2002–2017 and using time on study as time scale. The high and average PRS correspond to PRS values of 2 (mean + 2SD) and 0 (mean/median), respectively.

The plot indicates that the 10-year CVD risk for a non-smoking man with a high PRS (20.2%, 95% CI 17.0%–23.5%) exceeds that of a smoker with an average PRS (15.6%, 95% CI 14.1%–17.3%). Among women, a current smoker with an average PRS faces a comparable risk (8.4%, 95% CI 7.4%–9.5%) to a non-smoker with a high PRS (8.8%, 95% CI 7.6%–10.2%).

Communicating this information to healthcare professionals enhances their understanding of the importance of PRS and supports patient discussions, encouraging high-risk individuals to adjust their behaviour while considering the individual differences in genetic predisposition.

## DISCUSSION

To the best of our knowledge this is the first tailored model for a high CVD-risk population combining polygenic and traditional risk factors for estimating cardiovascular disease risk. This study found a significant improvement in risk discrimination for both men and women when incorporating the CAD PRS to the prediction model.

Our model used Estonian Biobank data and demonstrated a significant rise in the risk of CVD events and/or mortality with higher CAD PRS. Moreover, individuals with high PRS may experience the onset of the disease up to a decade earlier compared to those with medium or low PRS. This suggests that a high PRS (top 10%) constitutes a substantial risk factor, comparable to that of smoking or high cholesterol, with elevated risks apparent as early as in one’s 30s. Our study demonstrates that the PRS-inclusive risk model performed best in younger cohorts, which is crucial given the pressing need to develop high-quality risk assessment tools for younger populations, especially since existing models like SCORE-2 can only be used starting at the age of 40 [17]. Similar results have also been shown in previous PRS population-based models created with Finnish and UK Biobank data [6], [22], [33].

We constructed models utilizing two distinct cohorts from different time periods, yet the significance and consistency of the PRS effect remained consistent in both, underscoring its importance in clinical risk assessment, as also reported by Patel, et al [27]. The Estonian population is categorised as high risk for cardiovascular disease based on standardised cardiovascular disease mortality rates, thus serving as a proxy to multiple high-risk populations in Eastern Europe [17]. The baseline risk differs between the two Biobank cohorts, with the later cohort having lower levels of conventional risk factors resulting in lower CVD incidence rates compared to the earlier cohort (reflecting contemporary declining trends in CVD incidence in developed countries).Regardless of cohort differences, the inclusion of PRS in the model demonstrates its consistent effect on overall risk across both cohorts, highlighting the model’s robustness. As public health advancements and behavioural changes reduce the impact of traditional risk factors on disease risk, the relative importance of PRS grows [34]. Consequently, models like ours can help identify high-risk individuals within the population and guide targeted interventions.

The current European Society of Cardiology (ESC) Cardiovascular Disease Prevention Guidelines do not advocate for the routine collection of genetic data in primary prevention [32]. However, a recent clinical consensus highlights the critical need to quantify the potential benefits of PRS in clinical practice [35]. In European countries, polygenic risk scores are not yet integrated into routinely collected administrative health data for risk prediction at either the population or individual level. This practice should be reassessed as the global focus on prevention and health promotion increases, highlighting the growing importance of genetics and polygenic risk in everyday healthcare. Clinicians often rely on risk prediction models like SCORE2 and QRISK3 for cardiovascular risk management, but the potential of genetic testing in primary prevention requires more rigorous evaluation. Before integrating PRS into clinical practice, the effectiveness of PRS-based primary CVD prevention strategies, such as lipid-lowering treatments, must be validated through randomized clinical trials. As our study has revealed that individuals with very high CAD PRS face a significantly elevated risk of CVD, we argue that such individuals should be directed toward more intensive primary preventive measures at an early age [36].

### Strengths and limitations

A key strength of our study is the use of the Estonian Biobank database, which uniquely integrates clinical and genetic data—unlike most biobanks where these datasets are typically separate [37]. The availability of CAD PRS for nearly all biobank participants allowed us to leverage a large and comprehensive population for model development. This analysis was further enhanced by the high quality of the genetic data utilized to compute the polygenic scores as all SNPs were measured consistently using the same genotyping arrays.

Despite the EstBB cohort being a relatively large sample, a key limitation is the varied follow-up time for some individuals (7.3 years being the average follow-up time) and potential selection effects, due to volunteer-based sampling scheme. In addition, not all predictors had been uniformly measured for every individual, and the demographic composition, including ethnic and racial representation, reflects the Estonian population rather than Europe as a whole. To assure that the best possible model based on conventional predictors is used in both cohorts, accounting for possible differences from a random population-based sample, we did not rely on standardized risk-prediction algorithms such as SCORE2 but instead developed the models that provided the best fit for our data. To maintain simplicity, we chose not to include data on cardiovascular disease (CVD) treatment nor calculate the Charlson Comorbidity Index in the model [38].

## Conclusions

The results of this study emphasize the importance of using polygenic risk scores in combination with traditional risk factors for identifying individuals at high risk for atherosclerotic cardiovascular disease. From a primary prevention perspective, polygenic risk scores allow for the early assessment of risk, enabling the implementation of proactive prevention strategies aimed at reducing the burden of cardiovascular disease, particularly in a younger age.

## Acknowledgements

The Estonian Biobank Research Team, including Andres Metspalu, Lili Milani, Tõnu Esko, Reedik Mägi, Mait Metspalu, Mari Nelis, and Georgi Hudjashov, contributed to data collection, genotyping, QC, and imputation, while Priit Palta, Nele Taba, Erik Abner, Jaanika Kronberg, and Urmo Võsa contributed to the generation, development, and QC of the NMR data. The activities of the EstBB are regulated by the Human Genes Research Act, which was adopted in 2000 specifically for the operations of the EstBB. Individual level data analysis in the EstBB was carried out under ethical approval 1.1-12/624 from the Estonian Committee on Bioethics and Human Research (Estonian Ministry of Social Affairs), using data from the Estonian Biobank.

## Author contributions

K.F., L.M. and M.J. contributed to the conception of the study. T.P., K.F. contributed to the acquisition, analysis of the data included in the modelling. A.P and T.P. contributed to the interpretation of the data and drafted the manuscript. A.E., L.M., M.J. supported the interpretation of the data, and provided a scientific evaluation of the study making necessary adjustments.

## Data availability

The data sets supporting the findings of this study are available in aggregated format upon reasonable request from the corresponding author, ensuring compliance with confidentiality and ethical considerations.

## Supplementary Materials

**Table S1.** Endpoint definitions.

| **Fatal cardiovascular disease** |  |
| --- | --- |
| Hypertensive disease | I10–16 |
| Ischemic heart disease | I20–25 |
| Arrhythmias, heart failure | I46–52 |
| Cerebrovascular disease | I60–69 |
| Atherosclerosis/AAA | I70–73 |
| Sudden death and death within 24h of symptom onset | R96.0–96.1 |
| *Excluding* |  |
| Myocarditis, unspecified | I51.4 |
| Subarachnoid haemorrhage | I60 |
| Subdural haemorrhage | I62 |
| Cerebral aneurysm | I67.1 |
| Cerebral arteritis | I68.2 |
| Moyamoya | I67.5 |
| **Non-fatal cardiovascular disease** |  |
| Non-fatal ischemic heart disease | I21–25 |
| Non-fatal stroke | I60–69 |
| *Excluding* |  |
| Subarachnoid hemorrhage | I60 |
| Subdural hemorrhage | I62 |
| Cerebral aneurysm | I67.1 |
| Cerebral arteritis | I68.2 |
| Moyamoya | I67.5 |

## Management of predictors

All continuous predictors were centered and scaled before the analysis: age at 45 and by 5 years, BMI at 25 and by 5 kg/m2, SBP at 125 and by 20 mmHg, total cholesterol at 5 and by 1 mmol/L, HDL cholesterol at 1.5 and by 0.5 mmol/L. For BMI, values below 22 kg/m^2^ or above 40 kg/m^2^ were truncated at 22 and 40, respectively. For total cholesterol, values below 3 mmol/L or above 9 mmol/L were truncated at 3 and 9, respectively. Analogously, HDL cholesterol values were truncated at 0.7 mmol/L and 2.5 mmol/L. PRS for CAD was standardized to zero–mean and unit-variance.

**Table S2.**
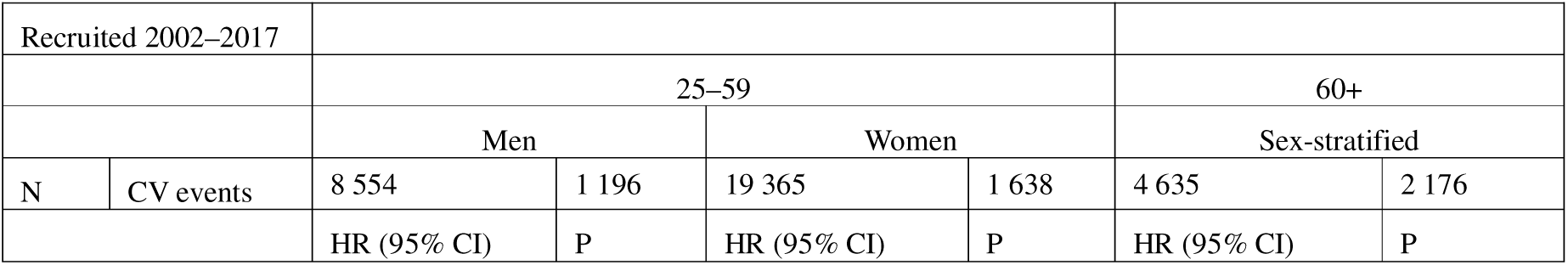
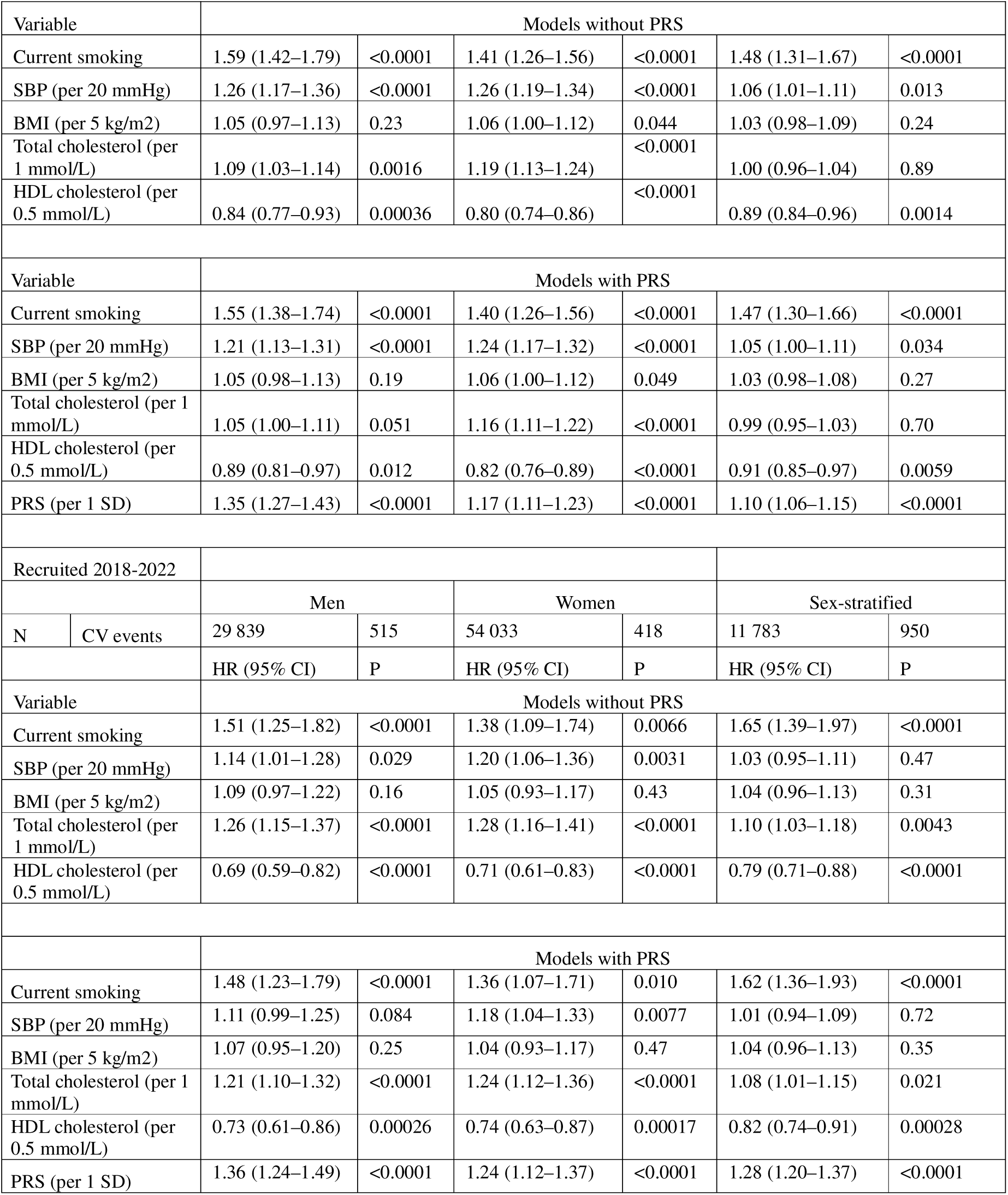
Hazard ratios and p-values from CVD risk models with and without the PRS. The models are derived in the full data of the earlier and later cohort using sex-specific and sex-stratified analysis for age groups 25–59 and 60+, respectively.

**Table S3.** Model discrimination in age and recruitment groups. The models are derived and the C-indices are calculated using the full data from the earlier and later cohort, with sex-specific and sex-stratified analyses for age groups 25–59 and 60+, respectively.

|  | Recruited 2002–2017 |  |  | Recruited 2018–2022 |  |  |
| --- | --- | --- | --- | --- | --- | --- |
|  | 25–59 |  | 60+ | 25–59 |  | 60+ |
|  | Men | Women | Sex-stratified | Men | Women | Sex-stratified |
| C-index of conventional model | 0.6029 | 0.6077 | 0.5438 | 0.6012 | 0.6146 | 0.5690 |
| C-index of conventional model with PRS | 0.6311 | 0.6124 | 0.5515 | 0.6258 | 0.6150 | 0.5997 |
| Difference in C-index | 0.0282<br>(P<0.0001) | 0.0047<br>(P=0.064) | 0.0077<br>(P=0.04) | 0.0246<br>(P=0.003) | 0.0004<br>(P=0.48) | 0.0307<br>(P=0.00048) |

## NRI analysis

Reclassification analyses comparing the conventional model to the conventional model with PRS were done separately for the two age groups (25–59 and 60+) but jointly for the two cohorts. Models were fitted separately for six subgroups based on cohort, age group, and sex. Specifically, there were separate models for the two cohorts, two age groups (25–59 and 60+), with separate models for men and women in the age group 25–59, and a sex-stratified model for the age group 60+. All models were developed using the training data. NRI was calculated according to predicted 5-year risk of CVD. The predictions were calculated for the individuals in the validation data. Individuals who died of other causes within 5 years were not included in these analyses.

Before calculating the NRI, the predictions for men and women in the 25–59 age group were combined, as were the predictions for the two cohorts, so that the NRI analysis included only two groups.

For the categorical NRI analyses, risk categories—low, intermediate, and high—were defined as <1.25%, 1.25% to 5%, and >5% within 5 years for the younger age group (25–59), and as <5%, 5% to 10%, and >10% within 5 years for the older age group (60+). The risk thresholds were slightly adjusted from the ones provided in the 2021 ESC Guidelines on cardiovascular disease prevention in clinical practice [1] to account for the different age groups used in this study and were halved to define 5-year risk thresholds based on the guideline-recommended 10-year CVD risk thresholds.

**Table S4.**
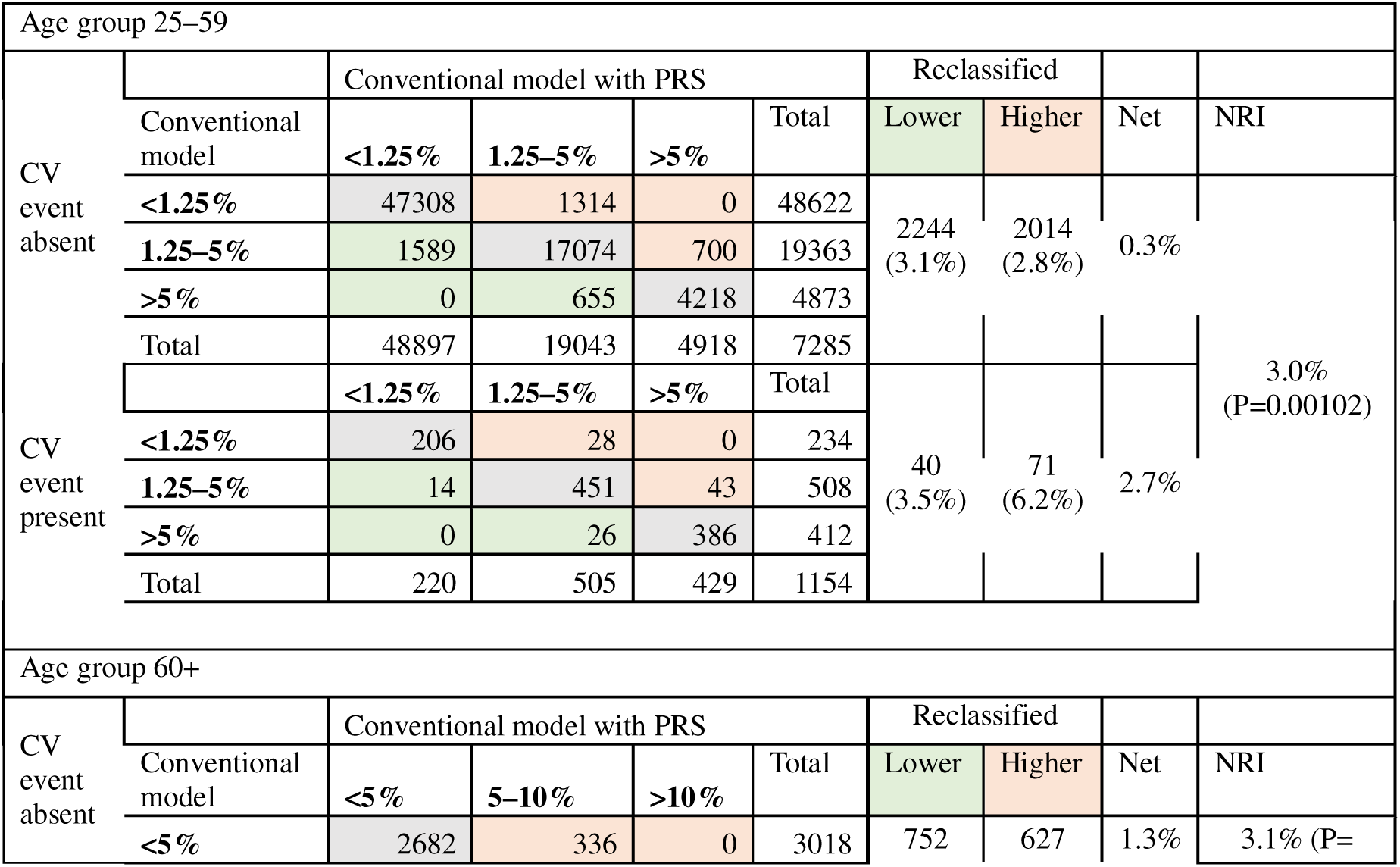
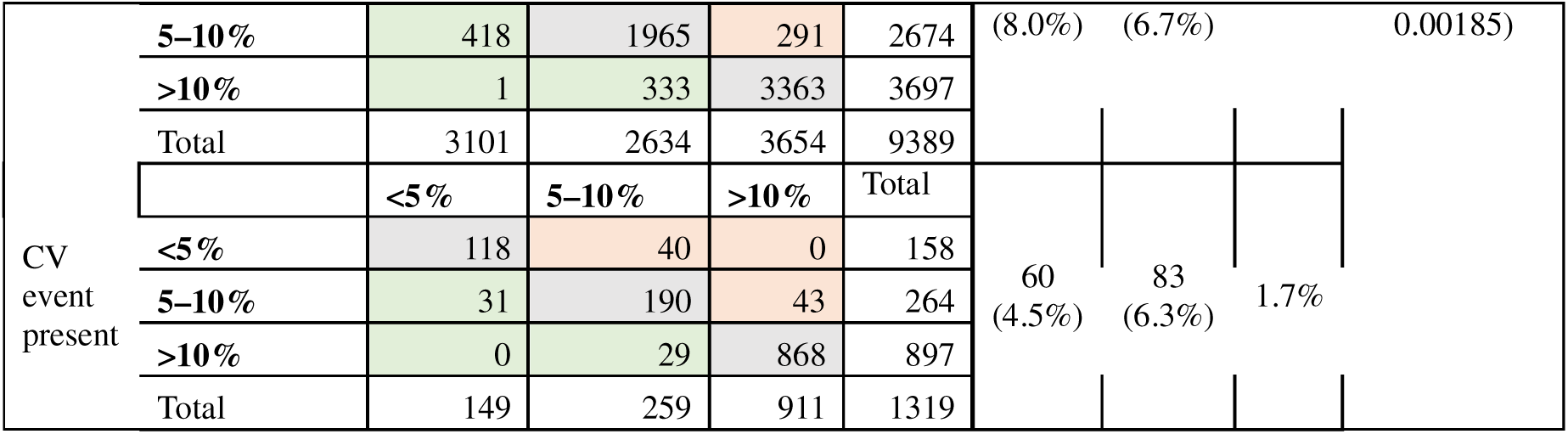
Categorical reclassification of 5-year CVD risk. The conventional model with and without PRS are compared separately in 25–59 and 60+ age group.

**Figure S1.**
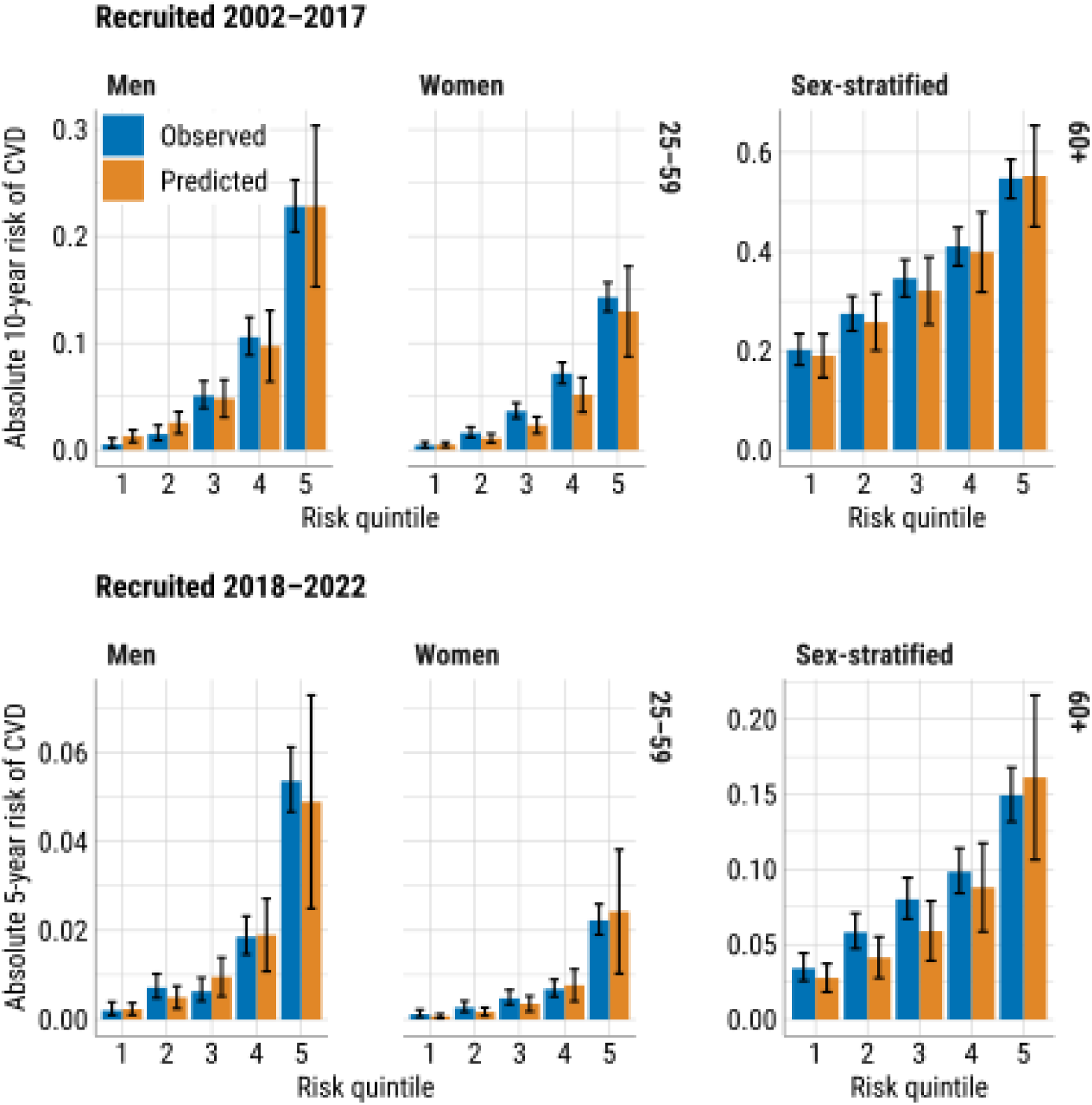
Calibration of absolute CVD risk in validation set. The calibration is assessed by sex, age group, and recruitment period. 10– and 5-year CVD risks are used for the earlier and the later cohort, respectively.

